# Exploring clinical characteristics of COVID-19 in children and adolescents using a machine-learning approach

**DOI:** 10.1101/2024.12.04.24318465

**Authors:** Stefania Fiandrino, Daniele Doná, Carlo Giaquinto, Piero Poletti, Micheal Davis Tira, Costanza Di Chiara, Daniela Paolotti

## Abstract

**Introduction:** The epidemiology and clinical characteristics of COVID-19 evolved due to new SARS-CoV-2 variants of concern (VOCs). The Omicron VOC’s higher transmissibility increased pediatric COVID-19 cases and hospital admissions. Most research during the Omicron period has focused on hospitalized cases, leaving a gap in understanding the disease’s evolution in community settings. This study targets children with mild to moderate COVID-19 during pre-Omicron and Omicron periods. It aims to identify patterns in COVID-19 morbidity by clustering individuals based on symptom similarities and duration of symptoms and develop a machine-learning tool to classify new cases into risk groups.

**Methods:** We propose a data-driven approach to explore changes in COVID-19 characteristics analyzing data collected within a pediatric cohort at the University Hospital of Padua. First, we apply an unsupervised machine-learning algorithm to cluster individuals into different groups. Second, we classify new patient risk groups using a Random-Forest classifier model based on sociodemographic information, pre-existing medical conditions, vaccination status, and the VOC as predictive variables. Third, we explore the key features influencing the classification.

**Results:** The unsupervised clustering identified three severity risk profile groups. The classification model effectively distinguished these groups, with age, gender, COVID-19 vaccination, VOC, and presence of comorbidities as top predictive features. A high number and longer duration of symptoms were associated with younger age groups, males, unvaccinated individuals, Omicron infections, and those with comorbidities. These results are consistent with evidence of severe COVID-19 in infants, older children with comorbidities, and unvaccinated children.

**Conclusion:** Our classification model has the potential to provide clinicians with insights into the children’s risk profile of COVID-19 using readily available data. This approach can support public health efforts by clarifying disease burden and improving patient care strategies. Furthermore, it underscores the importance of integrating risk classification models to monitor and manage infectious diseases.

## 1 Introduction

The epidemiology and clinical characteristics of COVID-19 evolved during the pandemic, largely due to the emergence of new SARS-CoV-2 variants of concern (VOCs) with different virulence and transmissibility. These changes in VOCs contributed to shifts in COVID-19 clinical manifestations and morbidity. The emergence of the B.1.1.529 (Omicron) VOC has been marked by a predominance of upper respiratory tract symptoms, such as rhinitis, cough, and sore throat, resulting in a lower incidence of severe outcomes among adults. However, the higher transmissibility of the Omicron VOC, combined with school reopenings [1], has led to a rise in pediatric COVID-19 cases [2–4], significantly increasing hospital admissions among children [5] and, consequently, severe outcomes in absolute terms.

Although the World Health Organization has declared an end to COVID-19 as a public health emergency [6], SARS-CoV-2 continues to persist and mutate. Coupled with a significant decline in global vaccination uptake and coverage, the risk remains of new VOCs emerging, potentially causing new surges in cases and deaths.

Given that the clinical characteristics of COVID-19 vary with different viral strains, understanding and early recognition of SARS-CoV-2 infection in the pediatric population is crucial to reducing the global burden of the pandemic [7, 8]. With the decline in testing, providing evidence on the clinical patterns of pediatric COVID-19 is essential for facilitating early recognition and prompt management of cases.

To date, most research describing the changing symptomatology of COVID-19 during the Omicron period has concentrated on hospitalized cases, focusing on more severe cases and limiting our understanding of the disease’s evolution in community settings, which represent the majority of cases [4, 9, 10].

This research focuses on the youngest population infected with mild to moderate COVID-19, covering pre-Omicron and Omicron infections from April 2020 to December 2022 in the Veneto region of Italy. The analyzed data consists of records of children aged 0-20 years seeking care from family pediatricians. The study aims to achieve two primary objectives: i) uncovering patterns in COVID-19 morbidity by clustering individuals according to the number and duration of symptoms experienced; ii) developing a machine-learning tool to classify new cases based on demographic data, treatments, and co-existing medical conditions, and vaccination status, using the classes of infection identified in the previous step.

This study builds on prior research by Di Chiara et al. [11], which investigated the epidemiological and clinical features of SARS-CoV-2 variants using descriptive statistics. Our research aims to reinforce these findings by employing an unsupervised machine-learning approach to analyze clinical manifestations in children. This approach helps clinicians to understand the classes of SARS-CoV-2 infections, thus the children’s risk profiles, and the possible burden of disease, facilitating better decision-making and personalized treatment [9, 12, 13].

## 2 Methods

### 2.1 Dataset description

In this study, we rely on data collected within a prospective cohort of 715 participants focusing on children and adolescents aged 0-21 years old attending the COVID-19 Family Cluster Follow-up Clinic (CovFC) from April 2020 to December 2022 [11]. The CovFC was instituted at the Department of Women’s and Children’s Health, University Hospital of Padua, situated in the Veneto region, Italy. Families, including children, older siblings, and parents, who had recovered from COVID-19 were referred to the CovFC by their family pediatricians (FPs), and to be eligible for the enrollment they had to meet two criteria: 1) have children under the age of 15, and 2) have one or more family members with a confirmed history of laboratory-confirmed COVID-19 infection. During enrollment, pediatricians and/or infectious diseases specialists conducted clinical assessments, including the collection of demographic information, medical history, SARS-CoV-2 virological test results from nasopharyngeal swabs, and vaccination status [14]. Clinical assessments and data collection were conducted for all individuals, including both parents and children, regardless of their laboratory-confirmed COVID-19 history. Following this, individuals with confirmed COVID-19 cases underwent a 6-monthly clinical and serological follow-up for at least one year after the initial infection, while subjects who were asymptomatic and had no analytical evidence of SARS-CoV-2 infection were considered non-COVID-19 cases. Vaccination data were recorded as they became available for each age group. Two blinded pediatricians determined the baseline infection date for each individual in the study, as outlined in [11].

In the current study, we implement additional specific exclusion criteria. Specifically, individuals classified as non-COVID-19 cases, and those older than twenty years were excluded from the analysis. Within the sample of 715 participants, 124 individuals were classified as non-COVID-19 cases, and 15 individuals were aged more than twenty years. Following the exclusion criteria, we discard those cases from the dataset, resulting in a final dataset including 581 children and adolescents.

### 2.2 Variables definition

We analyze data on existing medical conditions, vaccination status, and reported symptoms in the pediatric population, gathered through clinical assessments conducted at the enrollment. In terms of existing medical conditions, we first check the prevalence of each in the study sample, removing the ones without any representation in the dataset, and then we consider those among the list of comorbidities associated with severe pediatric COVID-19: chronic pulmonary conditions (e.g., bronchopulmonary dysplasia and uncontrolled asthma); cardiovascular conditions, (e.g., congenital heart disease); immunocompromising conditions (e.g., malignancy, primary immunodeficiency, and immunosuppression); neurologic conditions (e.g., epilepsy and select chromosomal/genetic conditions); prematurity; feeding tube dependence and other pre-existing technology dependence requirements; diabetes mellitus; obesity [15] [16] [17] [18]. To include vaccination status information, we deal with missing values reported in the dataset for the vaccination against COVID-19 due to the availability of the vaccines in the study period. For this reason, we consider the approval releases per age group: individuals older than 12 years old are considered vaccine-eligible from 31 May 2021 [19], while individuals aged 5-11 years old are vaccine-eligible from 01 December 2021 [20] and children younger than 4 years old were vaccine-ineligible when the enrollment was open. The individuals infected before the approval date of the vaccine were classified as non-vaccinated, and individuals aged 0-4 years old are all considered not vaccinated. Within the symptom set, non-referable symptoms for the younger age group, such as headache and small-taste alterations, have been excluded from the analysis. However, symptoms recognizable by parents, including myalgia and abdominal pain, have been retained. The final set includes fever, rhinitis, cough, dyspnea, myalgia, arthralgia, sore throat, conjunctivitis, asthenia, abdominal pain, nausea, lack of appetite, skin rash, confusion, ear pain, and other symptoms.

Starting from the available data, we extract additional information including the total number of symptoms reported during infection, the total number of comorbidities, the length of each symptom, the median duration of symptoms, the variant of infection, and the hexavalent vaccination (Diphtheria-tetanus-acellular pertussis, Polio, Hib, Hepatitis B). Specifically, we define an infection category for each individual, considering three types of infection: asymptomatic (duration of symptoms = 0 days), short (duration of symptoms *≤* 5 days), and long infection (duration of symptoms: *>* 5 days). These categories were defined in consultation with pediatricians who participated in the enrollment process. To identify specific variants of infections, we consider that from a clinical and immunovirological point of view, the Parental and Delta variants exhibited striking similarities. With the emergence of the Omicron variant, marked by substantial mutations in the S-RBD, a notable shift in the clinical, immunological, and epidemiological aspects of COVID-19 occurred. For these reasons, we classified cases into two groups based on the reported baseline date of infection onset: pre-Omicron and Omicron, defining any SARS-CoV-2 infection occurring before November 15, 2021, as pre-Omicron, and infections occurring after that day as Omicron. Finally, to include information on vaccination history, we combine available information on individual vaccinations and the hexavalent vaccination variable to determine whether an individual has received multiple vaccines intended to protect against several diseases (DTP, IPV/OPV, HBV, Hib).

### 2.3 Study Population

This study examines 581 children and adolescents who tested positive for SARS-CoV-2 (COVID-19 cases, symptomatic), aged 0-20 years old. The dataset includes socio-demographic and health-related information. The 66.5% of the study population (386 individuals) infected by SARS-CoV-2 were older than five years old, while the gender was balanced. Most of the subjects do not show previous underlying disease: only 23% of the entire study population exhibit at least one medical condition among the ones associated with severe pediatric COVID-19. During the pre-Omicron phase, almost all individuals haven’t done the COVID-19 vaccination yet, probably due to the vaccine-eligibility. At the same time, during the Omicron variant, the number of vaccinated children and adolescents increased (47 individuals out of 139 individuals infected during Omicron). As regards symptoms, only 7% of the infected people during the Omicron variant report no symptoms, while more than 65% present at least two symptoms. On the contrary, during the pre-Omicron period, nearly 35% of the individuals reported no symptoms, and less than 36% of the infected presented two or more symptoms. We provide a summary of clinical and sociodemographic characteristics (Table 1), including counts, percentages, medians, and interquartile ranges (IQR), as applicable. The stratification is based on the distinct phases considered, pre-Omicron and Omicron. We check the prevalence of each comorbidity and each symptom, to better characterize the cohort (Fig. 1 and Fig. 2). More than 75% do not exhibit any comorbidities, followed by individuals with other comorbidities, asthma, prematurity, and congenital heart disease (among the “others” category are all comorbidities not included in the listed ones). Lots of comorbidities included during the reporting phase do not show any representation in the dataset. For this reason, we remove them from the analysis, together with comorbidities not associated with severe COVID-19 in the pediatric age, that emerge to also have a low prevalence in the dataset (chronic hepatitis, rheumatic disease, nephropathy, hematological disease). We finally consider nine comorbidities: asthma, prematurity, congenital heart disease, neurological disease, diabetes, chronic respiratory disease, obesity, and others. The most common symptoms are fever and rhinitis, followed by headache, asthenia, and cough. As mentioned before, we do not consider both headache and smell and taste alterations for the analysis to avoid biases as they are non-referable symptoms for the youngest population. The most rare symptoms are confusion, polyadenopathy, and ear pain.

**Fig. 1.**
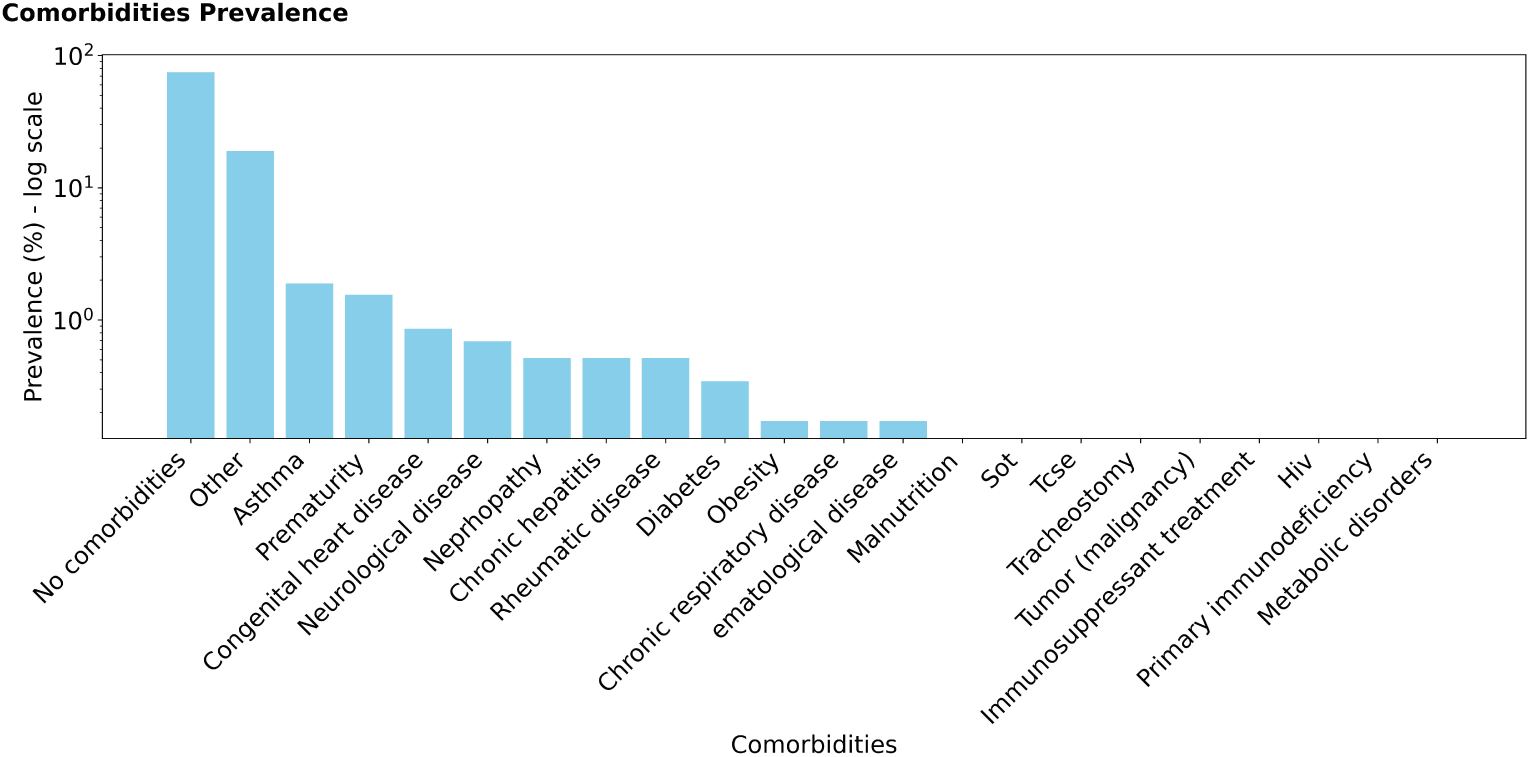
Characterization of study population: histogram of the prevalence of comorbidities in the dataset. A log-scaled y-axis has been used for readability.

**Fig. 2.**
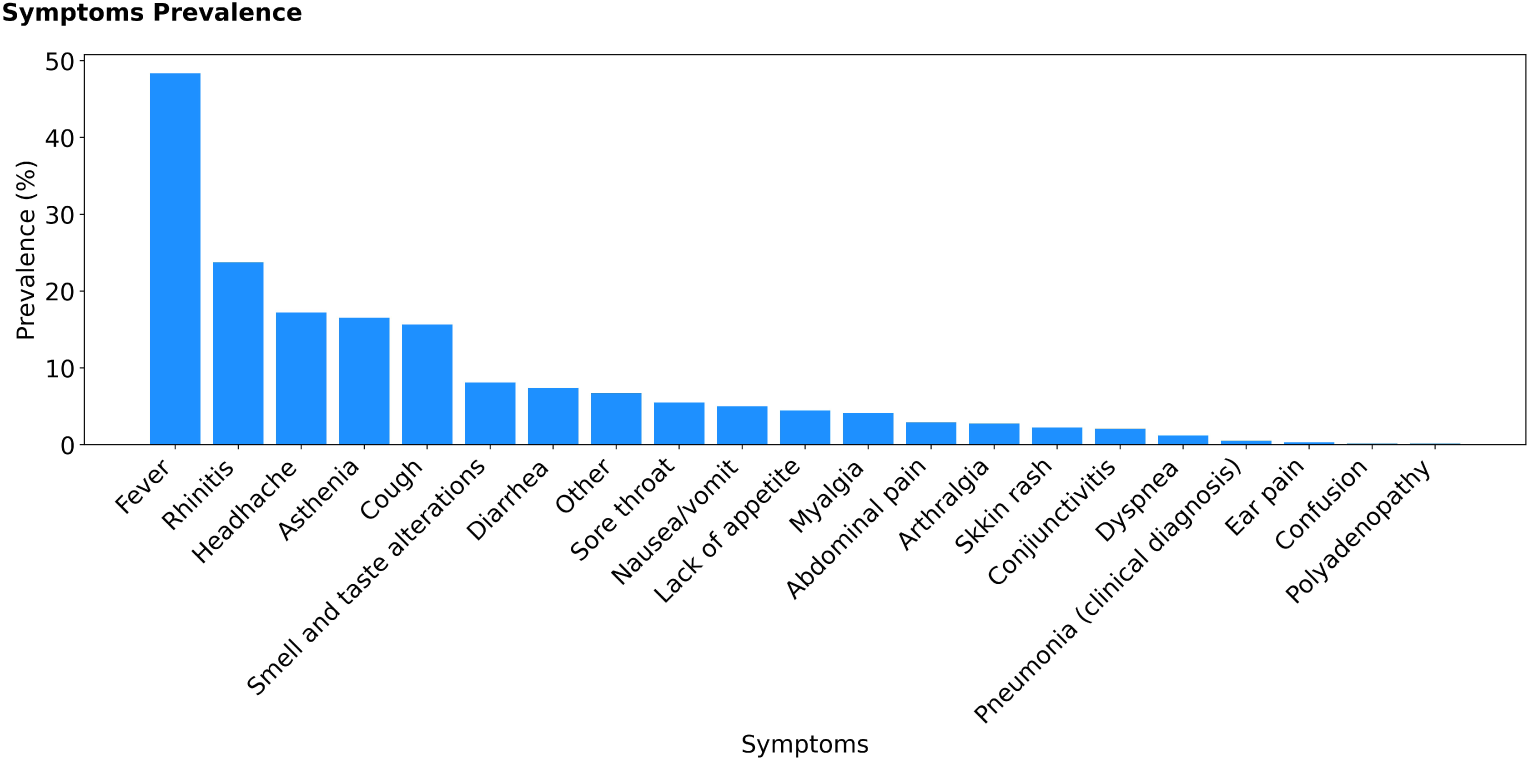
Characterization of study population: histogram of the prevalence of symptoms in the dataset.

**Table 1.**
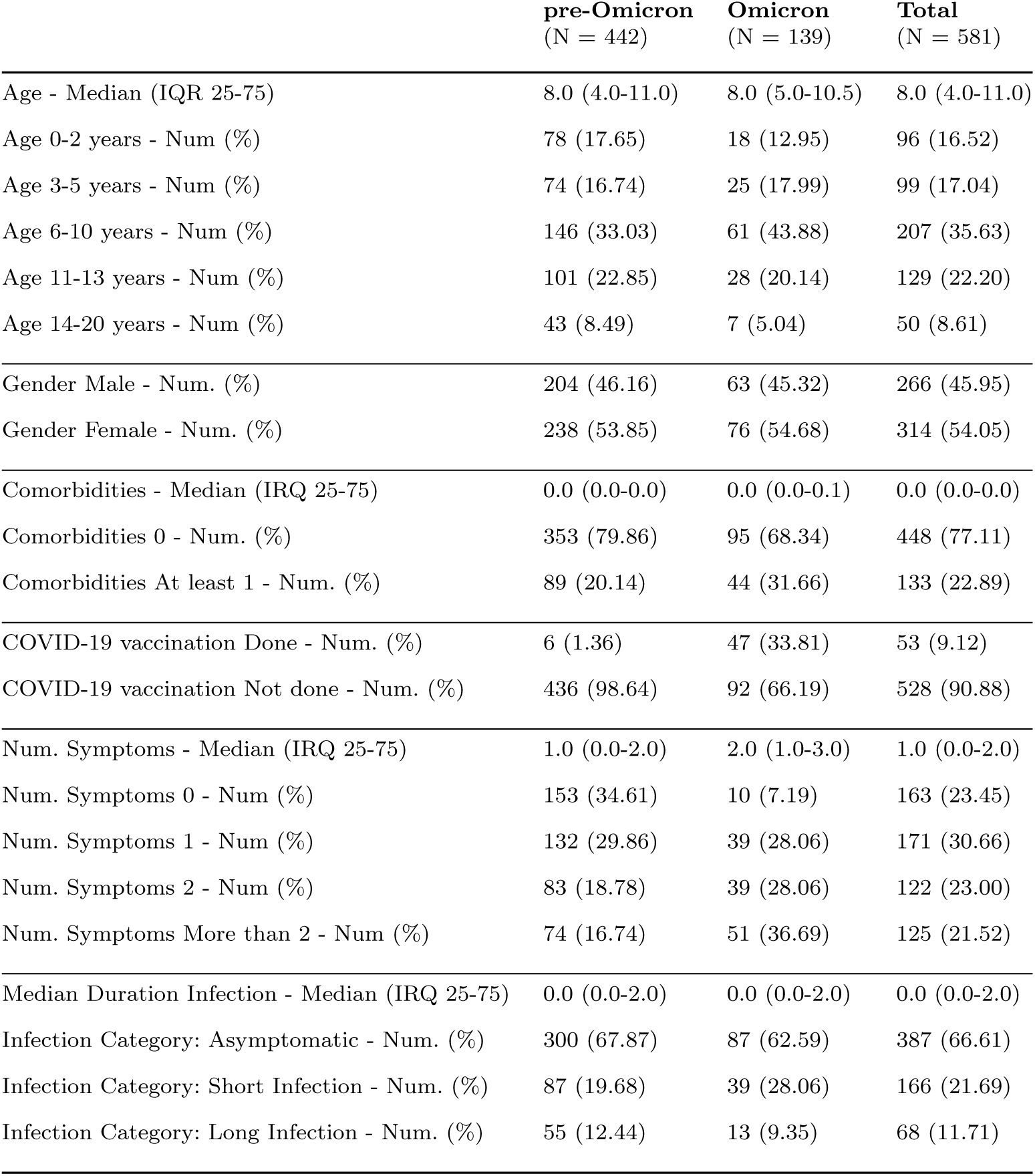
Overview of sociodemographic and clinical characteristics in the study population (N = 581), stratified by Omicron-infected and pre-Omicron group based on the SARS-CoV-2 variant.

|  | **pre-Omicron**<br>(N = 442) | **Omicron**<br>(N = 139) | **Total**<br>(N = 581) |
| --- | --- | --- | --- |
| Age - Median (IQR 25-75) | 8.0 (4.0-11.0) | 8.0 (5.0-10.5) | 8.0 (4.0-11.0) |
| Age 0-2 years - Num (%) | 78 (17.65) | 18 (12.95) | 96 (16.52) |
| Age 3-5 years - Num (%) | 74 (16.74) | 25 (17.99) | 99 (17.04) |
| Age 6-10 years - Num (%) | 146 (33.03) | 61 (43.88) | 207 (35.63) |
| Age 11-13 years - Num (%) | 101 (22.85) | 28 (20.14) | 129 (22.20) |
| Age 14-20 years - Num (%) | 43 (8.49) | 7 (5.04) | 50 (8.61) |
| Gender Male - Num. (%) | 204 (46.16) | 63 (45.32) | 266 (45.95) |
| Gender Female - Num. (%) | 238 (53.85) | 76 (54.68) | 314 (54.05) |
| Comorbidities - Median (IRQ 25-75) | 0.0 (0.0-0.0) | 0.0 (0.0-0.1) | 0.0 (0.0-0.0) |
| Comorbidities 0 - Num. (%) | 353 (79.86) | 95 (68.34) | 448 (77.11) |
| Comorbidities At least 1 - Num. (%) | 89 (20.14) | 44 (31.66) | 133 (22.89) |
| COVID-19 vaccination Done - Num. (%) | 6 (1.36) | 47 (33.81) | 53 (9.12) |
| COVID-19 vaccination Not done - Num. (%) | 436 (98.64) | 92 (66.19) | 528 (90.88) |
| Num. Symptoms - Median (IRQ 25-75) | 1.0 (0.0-2.0) | 2.0 (1.0-3.0) | 1.0 (0.0-2.0) |
| Num. Symptoms 0 - Num (%) | 153 (34.61) | 10 (7.19) | 163 (23.45) |
| Num. Symptoms 1 - Num (%) | 132 (29.86) | 39 (28.06) | 171 (30.66) |
| Num. Symptoms 2 - Num (%) | 83 (18.78) | 39 (28.06) | 122 (23.00) |
| Num. Symptoms More than 2 - Num (%) | 74 (16.74) | 51 (36.69) | 125 (21.52) |
| Median Duration Infection - Median (IRQ 25-75) | 0.0 (0.0-2.0) | 0.0 (0.0-2.0) | 0.0 (0.0-2.0) |
| Infection Category: Asymptomatic - Num. (%) | 300 (67.87) | 87 (62.59) | 387 (66.61) |
| Infection Category: Short Infection - Num. (%) | 87 (19.68) | 39 (28.06) | 166 (21.69) |
| Infection Category: Long Infection - Num. (%) | 55 (12.44) | 13 (9.35) | 68 (11.71) |

### 2.4 Unsupervised clustering

To uncover underlying patterns and structures within this dataset, we apply a clustering approach. Clustering is an unsupervised learning technique that categorizes data elements into groups based on inherent patterns, without requiring prior knowledge of the group definitions [21]. This method can be used to cluster the input data in classes based on their statistical properties. We aim to provide insights on the risk group of the SARS-CoV-2 infections in children based on the presence and/or absence of symptoms and the duration of the infection. The clustering output is a class label that characterize individuals based on the similarity of their reported symptoms (sixteen distinct types of symptoms represented as binary variables are involved), as well as the category related to the duration of the infection (three infection category are involved). We employ the K-modes algorithm [22], an extension of the well-established K-Means algorithm. K-means is well known for its efficiency in clustering large data sets. However, its limitation to numeric data restricts its applicability in fields such as data mining, where extensive categorical datasets are commonly encountered. Addressing the challenge of clustering large categorical datasets in data mining, Huang (1998) introduced the K-modes algorithm. This algorithm is a modification of the K-means and employs a simple matching dissimilarity measure tailored for categorical variables instead of the Euclidean distance. Unlike K-means, K-modes utilize modes instead of means for clusters and incorporate a frequency-based approach to update modes during the clustering process [23, 24]. The clustering procedure requires the definition of the number of groups to divide individuals, thus, to find the optimal number of clusters we implement the Elbow method. The Elbow method relies on the observation that as the number of clusters increases, the total cluster variance for a dataset decreases rapidly. However, when plotting total cluster variance against the number of clusters, beyond a certain point, this decrease slows down, resulting in a graph that resembles a bent elbow. The optimal number of clusters is determined by identifying the point at the elbow, where the decrease in cluster variance becomes stagnant [25]. More in detail, we consider different values for the number of clusters and compute the total within-cluster variance (WCSS) for each value. By plotting WCSS against the number of clusters, we look for a point where the graph sharply changes direction, forming an elbow. Beyond this point, the graph becomes almost parallel to the X-axis, indicating that adding more clusters yields diminishing returns in reducing variance. The *K* value at the elbow is considered the optimal number of clusters.

### 2.5 Classification model

The second research objective involves the development of a classification model to predict in which risk group a newly diagnosed individual should be assigned. This approach can help provide better-individualized treatments for COVID-19 patients in the future.

The risk groups are identified and defined by the output of the clustering. The predictive variables have been defined in consultation with pediatricians and include socio-demographic, vaccination status, comorbidities, and variant of infection information. In the following, we report the extensive list: age, gender, ethnicity, asthma, prematurity, obesity, diabetes, chronic respiratory disease, congenital heart disease, neurological disease, presence of at least one comorbidity, COVID-19 vaccination and hexavalent vaccination, pre-Omicron/Omicron period of infection.

We use the Random Forest classifier, a versatile and powerful supervised machinelearning algorithm. The Random Forest is an ensemble of tree-based classifiers, where each tree in the forest contributes a unit vote to predict the most probable class label for a given input [26]. This ensemble method is known for its speed, robustness to noise, and success in identifying non-linear patterns in data. Also, it can effectively handle both numerical and categorical data, and it is resistant to overfitting [27].

### 2.6 Explainability

The final goal of the analysis is to understand which are the features that drive the classification in different classes. Machine learning approaches are often perceived as black boxes, offering recommendations without revealing the underlying processes. Therefore, interpret the results, understanding the hidden patterns, and comprehend the reasoning behind the model’s conclusion play a key role, especially when model outputs are used to support decision-making. To interpret the prediction model’s output, we use the SHAP (SHapley Additive exPlanations) framework [28]. SHAP provides a unified measure of feature importance, aiming to understand each instance’s prediction by quantifying the contribution of each feature. Originating from cooperative game theory, the Shapley value addresses the issue of determining each player’s importance to the overall cooperation. Since features contribute to the model’s output as players with varying magnitudes and signs, Shapley values consider both the magnitude and direction of their contributions [29] and enable the visualization of the range and distribution of impacts on the model’s output [30].

## 3 Results

### 3.1 Characterization of clusters attributes

The clustering method aims to group individuals based on the similarity of types of symptoms and the infection category related to the duration of the symptoms. The Elbow method identifies three clusters as the optimal number, as shown in Fig. 3. Once obtained three groups of individuals, we perform statistical analyses to understand the underlying patterns, structures, similarities within the groups, and differences among them. The results reveal that the three clusters characterize individuals according to distinct levels of the total number of symptoms and median infection duration. Interestingly, this information was not used during the clustering process, but comes as a result and highlights the relevance of the machine-learning approach in distinguishing meaningful patterns in the data. The clusters can be characterized as follows: Cluster 0 represents individuals exhibiting few or no symptoms, suggesting a higher likelihood of asymptomatic infection; Cluster 1 and Cluster 2 include individuals with a higher number of symptoms; Cluster 2 includes COVID-19 cases with a longer likely duration of symptoms than people belonging to Cluster 1. Table 2 shows the descriptive statistics per cluster, together with the ANOVA one-way analysis to find which variables had a statistically different mean value between (at least two of those) the clusters. Cluster 1 and Cluster 2 differ in the similarity of reported symptoms, in particular for fever, rhinitis, and cough, and for the duration of the first two symptoms. Cluster 0 differs from the other two clusters because it captures the asymptomatic COVID-19 cases. Fig. 4 shows the histogram of the percentage of individuals per number of symtpoms. We find distinct patterns: within Cluster 0, there is a notable prevalence of individuals reporting no symptoms or a limited number of symptoms, while Cluster 1 and Cluster 2 show no representation among individuals reporting no symptoms; conversely, the behavior reverses for the occurrence of a high number of symptoms, where Cluster 1 and Cluster 2 are prominent, while Cluster 0 displays an opposing trend.

**Fig. 3.**
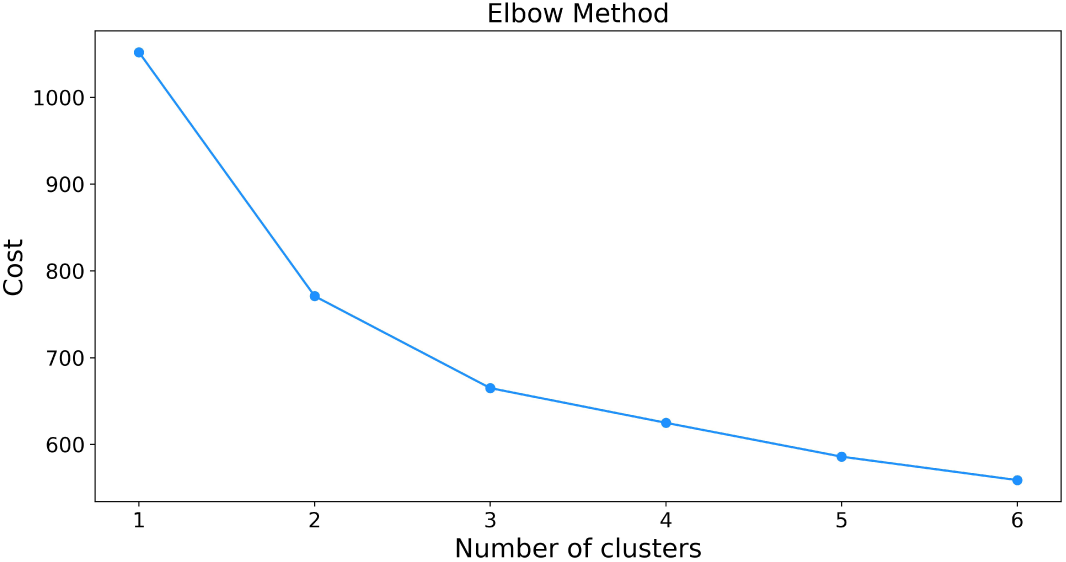
Clustering process settings: select the optimal number of clusters. A visual representation of the Elbow Method, in which the elbow point corresponds to three clusters.

**Fig. 4.**
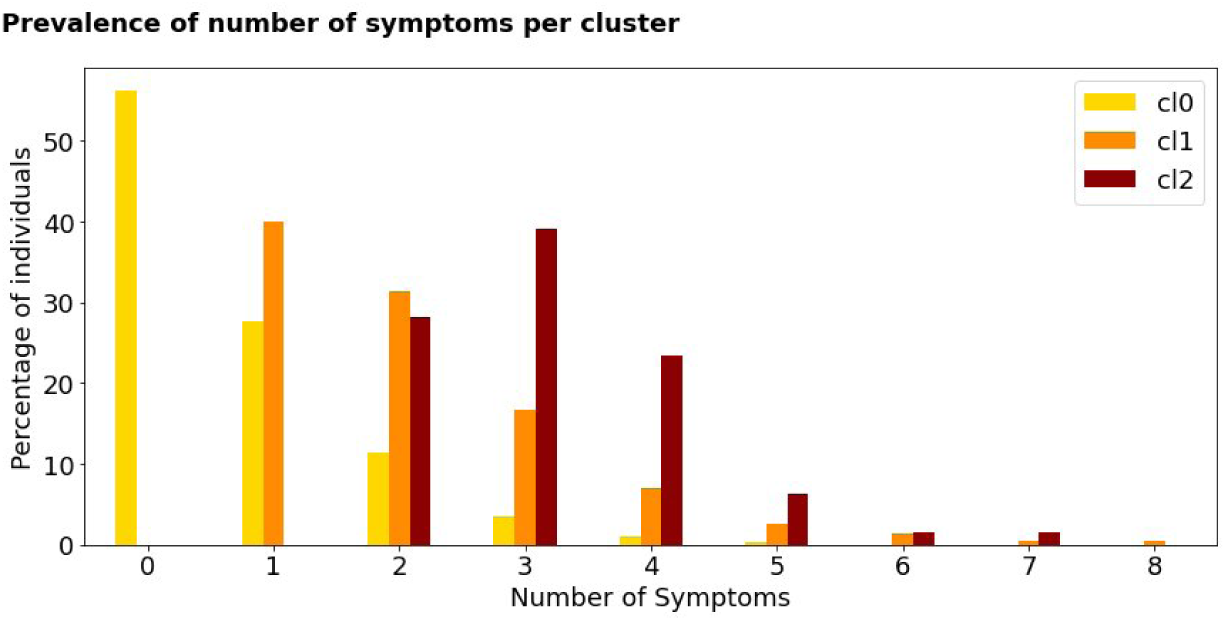
Clustering process results: histogram of the prevalence of the number of symptoms reported by individuals within different clusters. We observe different patterns: all the individuals reporting no symptoms are assigned to Cluster 0, while people reporting a higher number of symptoms belong to Cluster 1 and Cluster 2.

**Table 2.** Clustering process results: an overview of cluster characteristics in the study population (N = 581). Individuals are grouped based on the similarity of reported symptoms, and duration of symptoms category.

| Clinical characteristics | Cluster 0<br>(N = 290) | Cluster 1<br>(N = 227) | Cluster 2<br>(N = 64) | ANOVA<br>p-value |
| --- | --- | --- | --- | --- |
| Infection Category Asymptomatic (Num. (%)) | 225 (77.59) | 155 (68.28) | 7 (10.94) | 0.000 |
| Infection Category Short Infection (Num. (%)) | 42 (14.48) | 39 (17.18) | 45 (70.31) | 0.000 |
| Infection Category Long Infection (Num. (%)) | 23 (7.93) | 33 (14.54) | 12 (18.75) | 0.000 |
| Median Duration Infection (Mean (std.)) | 1.64 (8.38) | 2.71 (10.19) | 3.45 (2.78) | 0.219 |
| Number of symptoms (Mean (std.)) | 0.67 (0.92) | 2.09 (1.25) | 3.19 (1.07) | 0.000 |

### 3.2 Classification process

We use socio-demographic and clinical data to inform a Random Forest classifier and predict the risk group to which a new individual should be assigned. Given the dataset’s imbalance, we employed oversampling techniques using SMOTE to ensure reliable results [31]. In this work, 10-fold cross-validation is used to increase the models’ training effectiveness and lower the bias. Also, a grid search optimization approach is applied to choose the optimal parameters for the model, starting from a list of parameter alternatives.

The results of the model yield a Receiver Operating Characteristic (ROC) score of 0.73, indicating a 73% level of model performance in effectively distinguishing between the defined classes. Fig. 5 shows the confusion matrix, a visual representation of the actual versus predicted values, that measures the performance of the classification model. We report the raw confusion matrix and the row-wise normalized version, to better understand the percentage of correct classifications and errors across classes. The diagonal represents correctly classified instances, and off-diagonal elements represent misclassifications. Specifically, people with few or no symptoms (Cluster 0) were correctly classified for 55% of cases, and misclassified as belonging to Cluster 1 for 31% of cases. Individuals belonging to Cluster 1 were correctly classified for 48% of cases and misclassified as belonging to Cluster 0 for 32% of cases. Finally, COVID-19 cases in Cluster 2 were correctly classified for 72% of cases. Notably, when the model makes errors, it tends to misclassify individuals into the adjacent severity group rather than the more distant one. This pattern indicates that the model retains some discriminatory power, as it rarely assigns individuals from Cluster 0 directly to Cluster 2 or vice versa. Instead, misclassifications are more likely to occur between neighboring clusters, reflecting the severity levels. The cluster with the lowest correct classification rate is Cluster 1, representing a moderate level of symptoms. This cluster is the most challenging for the classifier, as it often misclassifies these individuals into extreme clusters (Cluster 0 or Cluster 2).

**Fig. 5.**
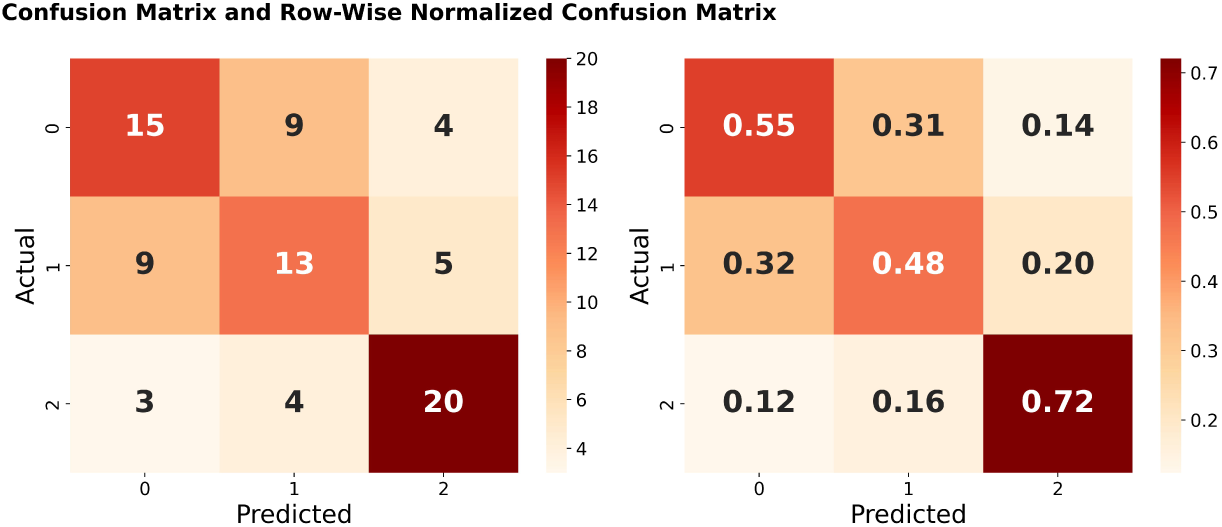
Multiclass classification model results: confusion matrices. On the left, the raw confusion matrix is shown, while on the right the row-wise normalized version.

### 3.3 Explanation of predicted values

To understand which predictive variables drive the Random Forest classifier outcome, we use SHAP (SHapley Additive exPlanation) values. Fig. 6A reports the SHAP summary plot, where features are first sorted by their global impact, and dots represent the shape values, colored by the value of that feature, from low (blue) to high (red). Age appears to be the most important factor, and the coloring shows a smooth decrease in the model’s output as age increases. Notably, we have similar results for gender, COVID-19 vaccination, and hexavalent vaccination, meaning that a lower risk profile characterizes females, and people with COVID-19 vaccination and hexavalent vaccination. On the contrary, a higher risk profile characterizes people infected during the Omicron variant, as shown by the opposite dot color distribution. Fig. 6B reports the mean absolute SHAP value of the features for the three classes, providing a general overview of the most influential features for the model (on the top) and their impact on the classification of each class. The top five predominant factors identified as crucial for the classification task are age, gender, the variant of concern (VoC), the presence of COVID-19 vaccination, and the presence of at least one comorbidity.

**Fig. 6.**
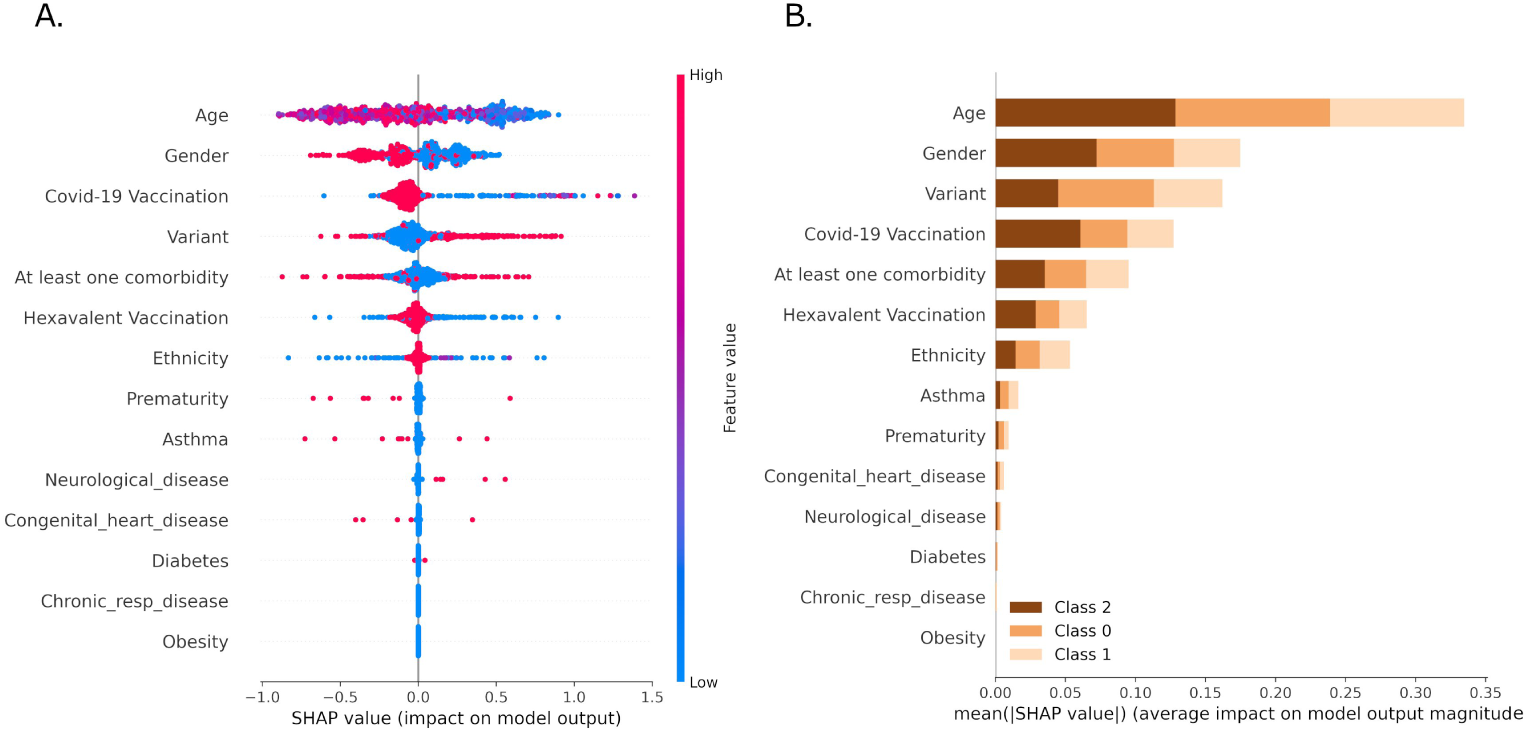
Model Explainability: a visual representation of the importance ranking of the risk factors with stability and interpretation: on the left (A), instance-individual SHAP values showing the impact on model output, with importance ranking of the top variables; on the right (B), global features importance based on the mean absolute magnitude of the SHAP values per class.

### 3.4 Discussion

The study presents a data-driven approach to exploring the characteristics of COVID-19 in children and adolescents during pre-Omicron and Omicron periods. We apply an unsupervised machine-learning approach to cluster individuals into different risk profile classes based on the similarity of types of symptoms and duration of symptoms category. Next, we classify the class in which a new patient should be assigned through a Random-Forest classifier model, using sociodemographic information, pre-existing medical conditions, vaccination status, and the VOC as predictive variables.

The unsupervised clustering approach identifies three risk profile groups, that result in different average numbers of symptoms and duration of symptoms: Cluster 1, characterizing individuals with fewer symptoms and most asymptomatic infections; Cluster 2, characterizing medium levels of number of symptoms and duration of symptoms; Cluster 3, including the most symptomatic cases.

Our analysis confirms the findings reported by Di Chiara et al. [11], where statistical and clinical descriptive approaches were employed. Clinical manifestations of COVID-19 in the youngest population vary according to the SARS-CoV-2 VOCs. Before the Omicron VOC emerged, 92% of identified infections in the community belonged to the lower risk groups (Cluster 0 and Cluster 1), with only 8% falling into the more severe symptom category (Cluster 2). However, during the Omicron period, this proportion increased to 22%, indicating higher symptomaticity in individuals seeking care, which may be partially explained by increased underreporting of SARS-CoV-2 infections during the brisk upsurge of Omicron cases.

We also examine specific sub-groups of symptoms including those that characterize an upper respiratory tract infection (i.e. cough, rhinitis, and sore throat). Among the Omicron-infected individuals, 58% of individuals showed at least one upper respiratory tract symptom, while among pre-Omicron infected 27%. This supports the evidence that Omicron infections seeking care were more likely to be linked to upper respiratory tract symptoms when compared to previous variants [2, 4, 11].

Irrespective of temporal changes in reporting rates, the classification model shows prominent and interesting results, being able to correctly classify 72% of the time individuals with more severe risk profiles, 55% individuals in the less severe group, and 48% individuals in the medium severe group. The model demonstrates its ability to distinguish between the three groups, with misclassifications typically occurring between similar levels of morbidity.

Age, gender, COVID-19 vaccination, VOC, and the presence of at least one comorbidity emerge as the top five features driving the classification process. Specifically, younger age groups among the age range (0-20 years old), male individuals, individuals without COVID-19 vaccination, individuals infected during the Omicron period, and individuals with at least one comorbidity tend to be associated with a higher risk profile group. In particular, focusing on the younger population (range 0-20 years old), COVID-19 symptoms still exhibit variation in clinical manifestations. Indeed, age emerged as a significant confounder driving the classification process. This aligns with previous evidence showing a higher risk of severe COVID-19 in infants, older children with comorbidities, and unvaccinated children [32–37].

### 3.5 Implications and applications

Despite COVID-19 becoming endemic among other seasonal respiratory viruses, the risk of new VOCs emerging with different virulence and transmissibility profiles, potentially leading to more severe cases, still persists. This work aids public health preparedness efforts and clinical decision-making. Furthermore, recent years have shown significant changes in the epidemiology and clinical presentation of seasonal respiratory viruses, with more severe cases of influenza and RSV among older children [38, 39]. In this context, a model that predicts infection type and progression based on a patient’s profile can guide clinical decisions, improving patient management and outcomes. As self-diagnosis becomes more common, it is crucial to recognize the limitations of self-diagnosis in terms of specificity and sensitivity, which can lead to misdiagnosis. Supplementing testing with clinical insights is essential to accurately identifying severity and risk profiles. The rise of self-testing also brings the risk of overtreatment, especially the overuse of antibiotics, which is a global health threat due to antibiotic resistance. A precise risk profile model can support clinicians in distinguishing between infections, helping to reduce unnecessary antibiotic prescriptions at the community level. Moreover, this model could be particularly beneficial in low- and middle-income countries where resources are limited. The ability to classify risk and predict disease progression using minimal resources can aid healthcare providers in these regions, improving patient outcomes.

### 3.6 Strenghts and limitations

Using data from a prospective cohort ensured more accurate and consistent data collection, limiting reporting bias. However, the present work comes with some limitations. The framework needs further testing on a substantially larger dataset, including the integration of socioeconomic information and the most severe cases such as hospitalized patients with the need for medical care (e.g., oxygen, ventilatory support). Similar to influenza, given the numerous variables that influence the risk of COVID-19, and the severity of the resulting illness, confounding is a significant issue in studies examining risk factors for COVID-19. Key potential confounders in these studies include socioeconomic variables such as household crowding, education level, and income [13]. Nonetheless, despite the limited size of our study population, the focus on mild and moderate cases, and the missing information on more detailed socio-economic aspects, we have identified differences in clinical manifestations among cases, highlighting distinct infection classes.

## 4 Conclusion

This data-driven approach provided different risk profile classes of COVID-19 in children using readily available information such as clinical history, VoC, vaccination status, and socio-demographic factors. This helps predict the risk profile group for a new patient. Overall, our findings highlight the importance of integrating risk-classification models to improve the management of infectious diseases, not only for COVID-19 but also for other respiratory infections. Further research is needed to profile classes of COVID-19 severity in children. This approach can support public health efforts by providing a clearer understanding of disease burden and facilitating better resource allocation and patient care strategies.

## Data Availability

Data are available from the corresponding author upon reasonable request.

## 5 Declarations

### 5.1 Ethics approval and consent to participate

The study protocol was approved by the local Ethics Committee (Prot. N° 0070714 of November 24th, 2020; last amendment Prot. N° 0024018 del 5/4/2022). Parents/legally authorized representatives were informed of the research proposal and provided written consent to participate in the study and use the collected patient data.

### 5.2 Consent for publication

Parents/legally authorized representatives were informed of the research proposal and provided written consent to use the collected patient data for research purposes and publication.

### 5.3 Availability of data and materials

Data are available from the corresponding author upon reasonable request.

### 5.6 Authors’ contributions

Dr. Stefania Fiandrino conceptualized and designed the study, performed the analysis, and wrote the manuscript. Dr. Daniele Donà performed the patients’ enrollment investigations, data curation, interpretation, and visualization, and contributed to the writing. Prof. Carlo Giaquinto conceptualized and supervised the study and contributed to the writing. Dr. Piero Poletti performed the validation, and methodology, and contributed to the writing. Dr. Michael Davis Tira performed the validation, and methodology, and contributed to the writing. Dr. Costanza Di Chiara conceptualized and designed the study, performed the patients’ enrollment and investigations, data curation and interpretation, supervised the study, and contributed to the writing. Dr. Daniela Paolotti conceptualized and designed the study, and methodology, supervised the study, and contributed to the writing. Dr. Costanza Di Chiara and Dr. Daniela Paolotti contributed equally as co-last authors. All authors had full access to all the data in the study, approved the final manuscript as submitted, and accepted responsibility for submitting it for publication.

## 5.7 Acknowledgments

The corresponding author would like to thank Dr. Bertilla Ranzato for her support in patient enrollment. The authors thank all the family pediatricians collaborating with the project. The authors thank all families who attended the COVID-19 family clusters follow-up Clinic of the University Hospital of Padova.

